# Bacterial Isolates and their Sensitivity Profile from CSF Samples – A 5 Year Study at a Tertiary Care Hospital

**DOI:** 10.1101/2023.03.24.23287676

**Authors:** Muhammad Moaaz Ali, Fatima Kaleem, Umme Farwa, Haider Ali, Saima Ishtiaq, Samina Javed, Saima Syed, Shahid Ahmad Abbasi

## Abstract

The death rates of medical situations like bacterial meningitis are always quite high. Cerebrospinal fluid (CSF) culture is the most common and reliable test to confirm the diagnosis of bacterial meningitis. All CSF samples that were received in the microbiology lab between January 2017 and December 2021 were included in the study. The study removed all duplicate samples and samples from patients who had recently started taking antibiotics. During the course of the trial, 2000 CSF samples were received. Just 157 of these samples successfully tested positive for the presence of infections. The results of the current investigation showed that the typical pathogens associated with meningitis were not isolated from CSF samples in our method. In contrast, in our setups, highly resistant microorganisms were isolated from CSF. primarily from patients with ventriculoperitoneal shunts.

## Introduction

Medical emergencies like bacterial meningitis always have significant mortality rates. The most common and effective inquiry to confirm the diagnosis of bacterial meningitis is cerebrospinal fluid (CSF) culture. The subarachnoid spaces of the cranium and spine, as well as the ventricles of the brain, are home to the ultra-filtrate of plasma known as cerebrospinal fluid (CSF). CSF helps to remove metabolic waste from the brain, including peroxidation products, glycolated proteins, extra neurotransmitters and debris from the ventricular lining, including bacteria, viruses, and other pointless substances. (1) A prevalent clinical infectious disease that is mostly brought on by bacteria entering the brain and spinal cord is central nervous system infection (CNS). Different microorganisms can infect CSF and cause illness. (2)Bacterial meningitis is the one of the serious diseases associated with substantial morbidity and mortality rates. The most common organism isolated from samples of bacterial meningitis are *Streptococcuspneumoniae, Neisseria meningitis* and *Haemophilusinfluenzae*. (3). Microbiology laboratories commonly receive CSF or blood specimens from patients with meningitis or unexplained febrile illness. Presumptive identification of *Neisseriameningitidis, S*.*pneumoniae*, and *H*.*influenzae* is sometime possible on the basis of cytological/microscopic examination of CSF.(4) Different methods exists for the diagnosis of bacterial meningitis of which CSF culture is still considered the gold standard.(5)CSF shunts are common neurological procedure performed for treatment of hydrocephalus. This treatment is often complicated by infection of shunt usually because of biofilm forming bacteria.. (6) Catheters used as CSF shunts are foreign bodies that can become infected with bacteria. The most common organisms in this case is Coagulase negative Staphylococci followed by Staphylococcus aureus. (7) Xentriculoperitoneal shunts infections are usually attributed to normal skin flora including the anaerobic microorganism like *Propionibacteriumacnes*. Gram negative bacteria and *candida* species are sometime also isolated. These organisms are usually thought to be introduced at the time of surgery. The incidence of shunt infections ranges from 10% -22% and its incidence is around 6% per procedure. Arround 90% of these infections occur usually in first 30 days after the procedure. The rate of these shunt infections is always influenced by some factors that are usually modifieable like hospital stay duration, experience of surgeon, duration and timing of procedure, procedure technique and indwelling device manipulation.*Neisseriameningitidis*, S. pneumoniae, *Hemophilusinfluenzae, E. coli*, and *K. pneumoniae* were shown to be the most frequently isolated bacterial etiologic agents from CSF cultures, according to recent investigations. The introduction of vaccines in the Extended Program of Immunization (EPI) against pneumococcai and *H. influenzae* type b has significantly reduced the burden of bacterial meningitis caused by these agents. In past 20 years the agents of disease, epidemiology and strategies of treatment for bacterial meningitis have change very significantly. The organisms like Streptococcus pneumoinae, Hemophilusinfluenza that were considered foremost important isolates for CSF infections but are now rarely isolated from CSF samples in our setup.Etiological pattern is changing so there is a need of finding out current pattern of isolates and their Antimicrobial profile from CSF samples. The changing The changing etiological pattern and changing antimicrobial profile needs serious attention, so as to give proper care to our patients.(8) At the same time, in our country there was no detailed study describing regarding CSF positive cultures. There are few reports regarding the resistance pattern of prevalent bacteria in this area. In order to provide useful information for developing strategies for preventing pathogens and enhancing evidence-based treatment, we thoroughly investigated antimicrobial resistance patterns and the common pathogenic bacteria of positive cerebrospinal fluid cultures in Tertiary Care Hospital Rawalpindi city, Punjab province from 2017–2021.

## Materials and Methods:-

This study was carried out at Department of Microbiology at Fauji Foundation Hospital Rawalpindi, Pakistan. This is a descriptive cross sectional study. Proper ethical approval was taken prior to start a study from Institutional Ethical FFH Review Board. All CSF samples received in microbiology Lab during the duration of study (Jan 2017-Dec 2021) were included in the study. All duplicate samples and samples of patients already on antibiotics were excluded from study. A total of 2000 CSF samples were received during the study during the study duration. Out of these samples around 18 duplicate samples were excluded from study.

CSF routine examination was also done on all samples.ThereceivedCSFsamples were centrifuged at 2500 r/min in the laboratory of Microbiology. The supernatant fluid was discarded. The sediment was inoculated on blood agar, Chocolate agar and MacConkey’s agar and were incubated at 370C for 24-72 hours aerobically and in 5 % CO2.

Cultures that yielding growth of any bacterial agent were further proceeded by standard microbiological methods of bacterial identification like Gram Stain, catalase, Coagulase API 20E and API 20NE (Biomeurix). All the isolated pathogens were identified at the species level. All microorganisms were subjected to the antimicrobial sensitivity testing by modified Kirby Bauer Disc diffusion method on Muller hinton agar (Oxoid). The isolates were incubated with antimicrobials for 18-24 hours at 37^0^C. The applied antimicrobials disks along with their strengths are as follows: Amikacin 30μg, Ampicillin 10μg, Augmentin 30μg, Azithromycin, Ceftazaidime, Ceftriaxone 30μg, Chloremphenicol 30μg, Ciprofloxicin 5μg, Colistin, Cortimoxazle 25μg, Doxicycline30μg, Gentamycin 10μg, Imepenem 10μg, Cloxacilin, Liniziolid 20μg, Methicillian 30μg, Erythromicin 15μg, Augmentin, Meropenem 10μg, Cephradine 10μg, Chloramphenicol 30μg, Penicilin 10μg, Vancomycin 30μg, Cotrimoxazle 25μg. The results of the antimicrobial sensitivity were interpreted according to the Clinical Laboratory Standards institute Criteria (CLSI 2021) (9). Dependent variable like frequency of bacterial meningitis causing isolates pattern of their antimicrobial resistance and Independent variables like socio demographic data of patients were calculated using the SPSS ver 21.

## Results

A total of 2000 CSF samples were received during the study period. From these samples only 157 samples yeailded the growth of pathogens. Over the period of 5 years the culture positive ratio has increased significantly as depicted in the Fig 1.

**Figure 1.**
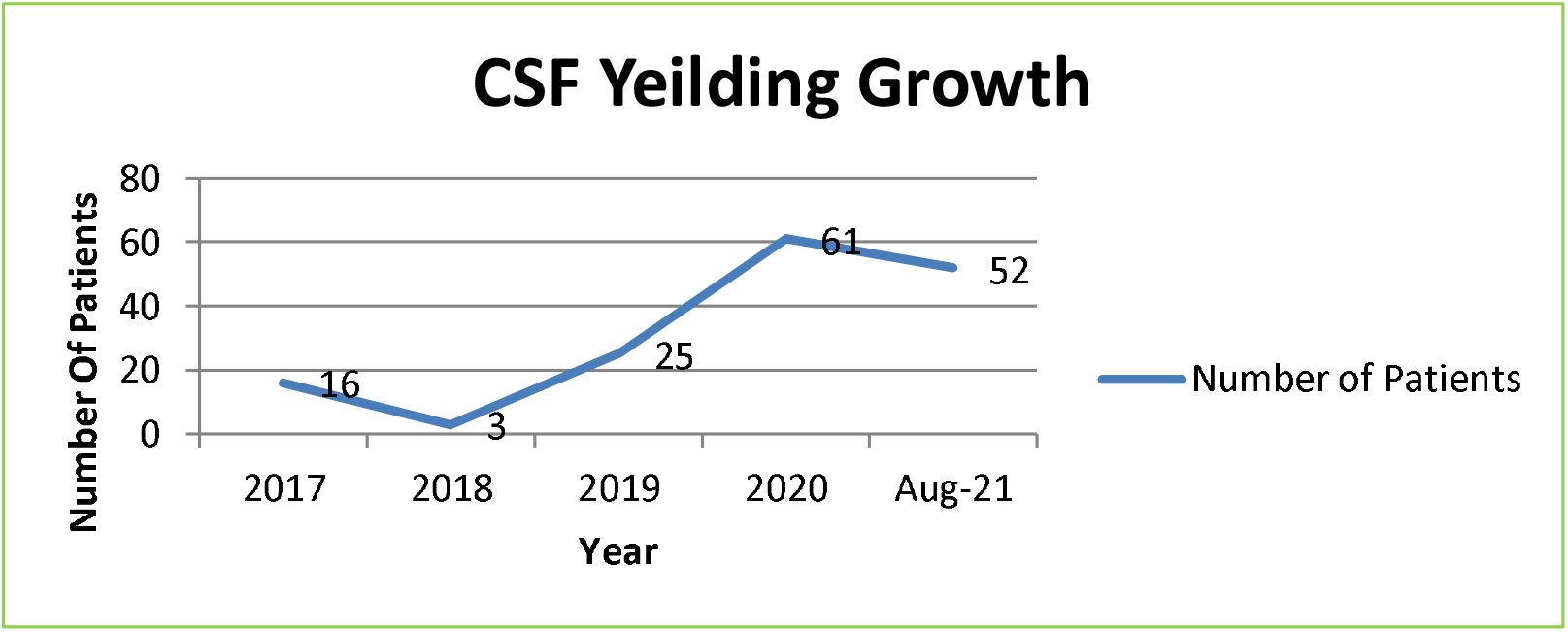

The ratio of positive cases in female neuro surgery ward was much higher from rest of the wards/departments of the Hospital. Fig 2.)

**Figure 2.**
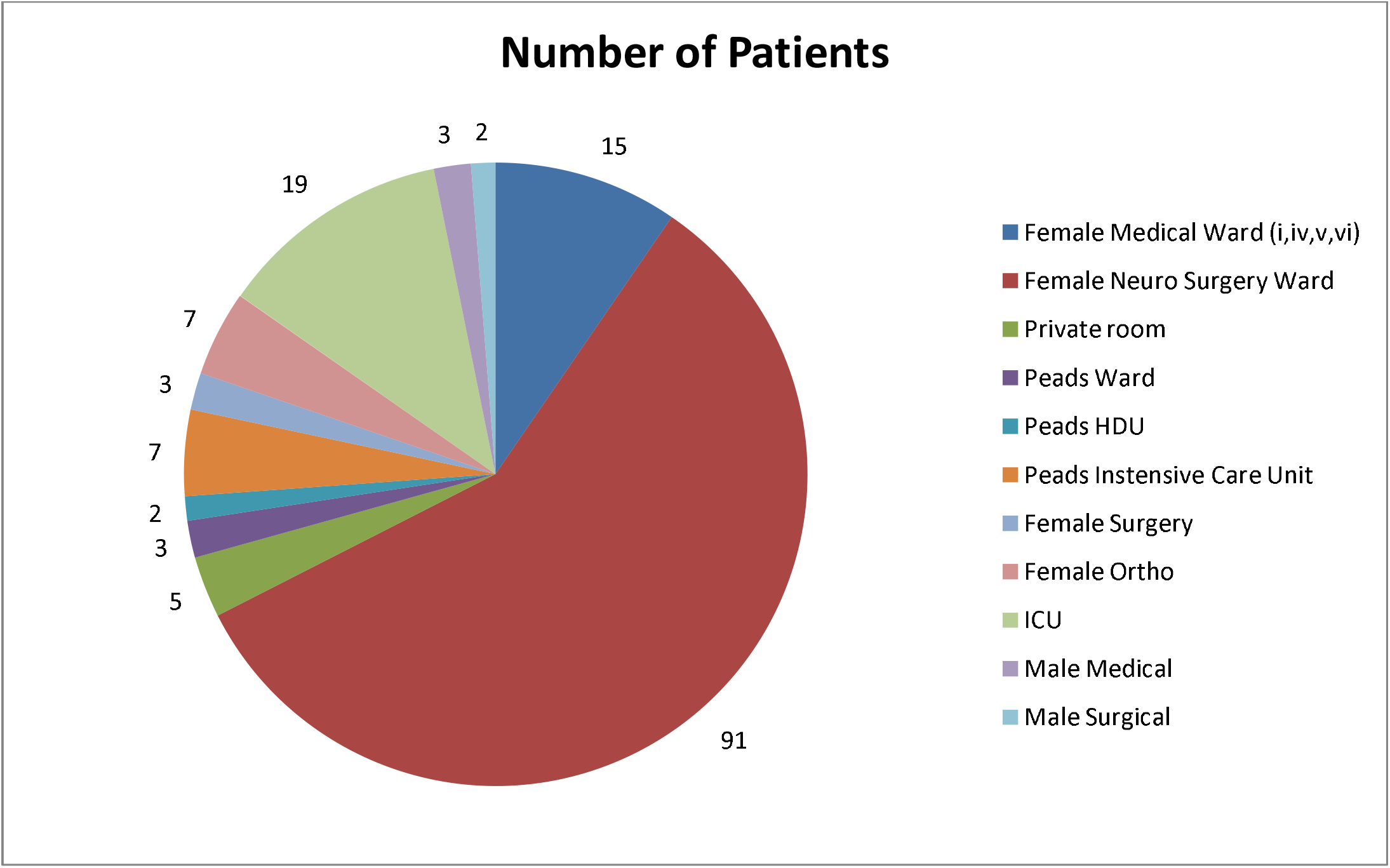

### Drug sensitivity Analysis of Gram Negative Pathogens in Cerebro Spinal fluid(Table 1)

In cerebrospinal fluid culture positive specimens, Escherichia coli (15.9%), Enterococcus species (10%), Klebsiellapneumoniae (14.6%), and Pseudomonas species (30%) were the most common bacteria isolated. Methecillin Resistant Staphylococcus Epidermidis (1.2%), CONS (1%), Methecillin Resistant Staphylococcus Aureus (11.4%), Staphylococcus aureus (3.1%), Patients under 25 years old had the majority of positive cultures. G-ve bacteria differ significantly by gender, age, and season; Escherichia coli was more prevalent in people under the age of 15 and Pseudomonas species were more prevalent in those under the age of 25. Summer was the season with the highest percentage of positive cultures.An interesting case of a female girl, 20 years old having CSF Shunt, was also included in study, Her first culture showed positive CSF culture growth of *E, coli, Pseudomonas* and *Enterococcusfaecalis*. After long hospital stayanduse of antibiotics the candida species were also isolated later in CSF of the patient.

**(Table 1).**

| Drugs | *E. Coli*<br>n=25(1<br>5.9%) | *Klebseila*<br>Oxytoca<br>n=2(1.2%) | *KlebseilaPne*<br>*umoniae*<br>n=23(14.6%) | *Pseudomonas*<br>*aeruginosa*<br>n=34(21.6%) | *Acinatobacter*<br>*baumannii*<br>n=19(12.1%) | *Pseudomona*<br>*s Species*<br>n= 13(8.2%) |
| --- | --- | --- | --- | --- | --- | --- |
| Amikacin30μ | 7(28%) | 1(50%) | 6(26%) | 18(53%) | 2(10.5%) | 7(53%) |
| g |  |  |  |  |  |  |
| Ampicillin<br>10µg | Nil | Nil | Nil | Nil | 2(10.5%) | Nil |
| Augmentin<br>30µg | 1(4%) | Nil | Nil | Nil | Nil | Nil |
| Azithromycin<br>15µg | Nil | Nil | Nil | Nil | Nil | Nil |
| Ceftazidime<br>30µg | Nil | Nil | Nil | 10(29%) | Nil | 3(23%) |
| Ceftriaxone<br>30µg | 1(4%) | Nil | Nil | 5(14.7%) | 1(5.2%) | 1(7.6%) |
| Chloremphenicol<br>30µg | 7(28%) | Nil | Nil | Nil | 3(15.7%) | Nil |
| Ciprofloxacin<br>5µg | 2(8%) | Nil | 4(17.3%) | 26(76.4%) | 5(26%) | 10(76%) |
| Colistin | Nil | Nil | 18(78.2%) | Nil | Nil | Nil |
| 300µg |  |  |  |  |  |  |
| Cortimoxazole<br>25µg | 1(4%) | Nil | Nil | Nil | Nil | Nil |
| Doxycycline<br>30µg | 5(20%) | Nil | 1(4.3%) | Nil | 5(26%) | Nil |
| Gentamycin<br>10µg | 6(24%) | Nil | 4(17.3%) | 21(61%) | 1(15.2%) | 7(53.8%) |
| Imepenem<br>10µg | 9(36%) | Nil | 8(34.7%) | 34(100%) | 5(26%) | 8(61.5%) |
| Meropenem<br>µg | 6(24%) | 1(50%) | 8(34.7%) | 25(73%) | Nil | 7(53.8%) |
| Minocycline<br>30µg | 5(20%) | 1(50%) | 3(13%) | Nil | 1(5.2%) | Nil |
| Polymixin B<br>300µg | 10(40%)<br>) | 1(50%) | 9(39%) | 28(82.3%) | 6(28.5%) | 12(92.3%) |
| Sulzone µg | 2(8%) | 1(50%) | Nil | 19(55.8%) | Nil | 7(53.8%) |
| TigeCycline<br>µg | 18(72%)<br>) | Nil | 4(17.3%) | Nil | Nil | Nil |
| Cefotaxime<br>105µg | Nil | Nil | Nil | Nil | Nil | Nil |

### Drug sensitivity Analysis of Gram Positive Pathogens in Cerebro Spinal fluid (Table 2)

**(Table 2).**

| Drugs | *MRSA*<br>n=20(12.5%) | *Staphylococusaureus*<br>n=5(3.1%) | *Enterococcusfaecalis*<br>n=16(10%) |
| --- | --- | --- | --- |
| Cephhradine 10µg | 1(5%) | 2(40%) | Nil |
| Chloramphenicol<br>30µg | 10(50%) | 4(80%) | 9(56.25%) |
| Ciprofloxacin 5µg | 5(25%) | 3(60%) | 4(25%) |
| Doxycycline 30µg | 16(80%) | Nil | 13(81.25%) |
| Gentamicin 10µg | 15(75%) | 3(60%) | 1(6.25%) |
| Penicillin 10µg | 5(25%) | 3(60%) |  |
| Vancomycin 30µg | 17(85%) | 5(100%) | - |
| Cotrimoxazole 25µg | 8(40%) | 3(60%) | - |
| Methicillin 30µg | Nil | 5(100%) | - |
| Amikacin 30µg | Nil | - | - |
| Ampicillin 10µg | Nil | - | 6(37.5%) |
| Ceftriaxone 30µg | 12(60%) | - | Nil |
| Cloxacilin µg | 8(40%) | 2(40%) | Nil |
| Lineziolid 20µg | 16(80%) | 4(80%) | 10(62.5%) |
| Imepenem 10µg | Nil | Nil | Nil |
| Meropenum 10µg | Nil | Nil | 5(31.25%) |
| Erythromycin 15µg | Nil | Nil | 2(12.5%) |
| Augmentin 30µg | Nil | Nil | 5(31.25%) |

## Discussion

One of the most fatal illnesses is bacterial meningitis. Even while effective antimicrobials and immunizations have decreased incidence and improved patient care, the mortality rate can still reach 34% in developing countries like us. Meningitis causes severe side effects like brain damage, hearing loss, long-term consequences like epilepsy, and mental instability etc. All these complications are more common if bacterial meningitis is acquired during early child hood.Despite the availability of many broad spectrum antibiotics there is high level of resistance in pathogens causing CSF infections. The mortality associated with these CSF infections ranges from 16-32% (12-16). The incidence of these infections after surgery i.e 8% still remains very high in developing world. The frequency of CSF culture positivity was constantly high in our setup. It is very necessary to analyze the epidemiology and drug resistance of CSF microbial infection.Despite improvements in the epidemiology of bacteremia in this population, the prevalence of group B Streptococcus remains the principal cause of meningitis (18). In 90% of cases of meningitis, bacteria have been found to be the cause (Strep pneumonia). 86% of cases were brought on by H. influenzae, 75% by N. meningitides, and 50% by Gram-negative bacilli. (19). Due to miss use of antibiotics in our country we analyze that the documented pathogens which were present in positive cultures of over setup were not seen in our hospital. The second main observation in our study was that the patients of shunt showed more growth of these organisms in our setup. On the other hand the resistant pattern of antibiotics in our setup werelittle more differing from our neighboring countries as indicated by studies come out from India and Iran(20)Our study identified *Pseudomonasaeruginosa*, it was the most prevalent microorganism and it was identified in 21.6% of culture in this study.Klebsiellapneumoniae is also seen very commonly isolated from CSF samples in patients with critical medical conditions, leading to high mortality rates (33-48%) (21-22)the timely detection of such pathogens and having a consideration for them in empirical therapy may improve critical outcomes. (23).The main issue in Klebisiella isolates is the emerging resistance against beta lactams and even carbapaenems. (24).A meta analysis study carried out in Pakistan revealed that pathogenic E.coli and Klebsiellaspecies isolated from CSF samples and other important samples are highly resistant organisms and they were resistant to Penicillins 100%, ampicillin 91%, amoxicillin 85%, ceftaxime 82% and cefaclor 100%. (25) Wefound*MRSA*and*Staph aureus*arehighlyresistanttomostofantibiotics. Similarly in various reports from Pakistan reveal high resistance of Staphylococcus aures to Penicillins (98%), Cefoxitin and other antimicrobials. (26). This high resistance level is due to easy availability of antimicrobials, self mdications and taking incomplete dosage of antimicrobials. (27)

## Conclusions:-

The present study identified that, the common organisms thought to be causative factor of meningitis were not isolated from CSF samples in our setup. Instead highly resistant pathogens were isolated from CSF in our set-ups. Mostly from patients having ventriculoperitoneal shunts.

## Recommendations:-

In our country, additional studies are recommended of all genders, is highly needed enhancing the surveillance and timely reporting of antibioticresistance is urgently needed in Pakistan.

### Limitations of the Study:-

Our research has potential limits. This hospital is only available for Hospital Staff, retired military person’s families and their children. So the ratio of patients of woman’s is average higher than men’s.

So study will be one sided the positive ratio of children and females is much higher thanmen’s. We have collected 5 years data. The sample size is small, which may affect the results and produce deviations.

## Data Availability

All data produced in the present study are available upon reasonable request to the authors
All data produced in the present work are contained in the manuscript
All data produced are available online at

## Notes

### Competing Interest Statement

The authors have declared no competing interest.

### Funding Statement

This study did not receive any funding

### Author Declarations

Proper ethical approval was taken prior to start a study from Institutional Ethical FFH Review Board.

## References

1. Sakka L, Coll G, Chazal J. Eur Ann Otorhinolaryngol Head Neck Dis. 2011 Dec;128(6):309–16. doi: 10.1016/j.anorl.2011.03.002. Epub 2011 Nov 18.

2. Teng Z, Mengjiao Kuong S, Havang Jizhen Li, QipengXie. Department of Lab Medicine. Children hospital Wenzohu:Zhejiang; 2021 June3.

3. Walaa SK, Safia HE. Identificationofcommonbacterialpathogen:doi: 10.1155/2016/4197187 PMID: 27563310 2016 August 01.

4. WHO meningitis Manual Author: 2011.. https://apps.who.int/iris/handle/10665/70765

5. David M, Firm, Maureen L, Schmidekand Sweetoperative Neurological Techanique:doi:

6. Alia Hdeib, Alan RCP Noleakgical Sugay Nouoolbgical 13rd edition) DOI: 10.1111/epi.124472012.10.1043/1079-0268 (2006) 29[240:SASONT]2.0.CO;2 6thedition;2012sep5.

7. Jayaraman Y, veeraraghavan B, chethrapilly, GK et al. Burden of bacterial meningitis in India. Preliminary data from a hospital based sertinalservieillance network. PLOS one 2018 13(5) e0197198.

8. Demissie Assegu. Fenta KL, Barrie D. Antimicrobialsensitivity Hawasa: University HOSPITAL; 19 May 2020. DOI:10.1186/s12866-020-01808-5

9. CLSI. Performance Standards for Antimicrobial Susceptibility Testing. DOI https://doi.org/10.1128/AAC.02631-20 30th ed. 2021. CLSI supplement M100-ED30(16)

10. Hum VaccinImmunother. 2018; 14(9): 2142–2149. 2018 Jul 9. doi: 10.1080/21645515.2018.1476814 PMID: 29787323

11. Philippe B,Sibe A, Yves B, Impact of vaccines on antimicrobial resistance, Doi https://doi.org/10.1016/j.ijid.2019.10.005, mOctober 7 2019

12. Jarousha AM, Afifi AA. Epidemiology and risk factors associated with developing bacterial meningitisamongchildren in gazastrip. Iran JPublicHealth. 2014;43(9):1176–1183. DOI 10.1186/s12887-015-0416-6

13. Shrestha RG, Tandukar S, Ansari S, et al. Bacterial meningitis in children under 15 years of age in Nepal. BMCP ediatr. 2015; 15 (1):94. doi:10.1186/s12887-015-0416-6

14. McCormick DW, Wilson ML, Mankhambo L, et al. Risk factors for death and severe sequelae in Malawianchildrenwithbacterialmeningitis, 1997–2010. Pediatr Infect Dis J. 2013;32(2):e54.–61. doi:10.1097/INF.0b013e31826faf5a

15. Namani S, Milenković Z, Kuchar E, et al. Mortality from bacterial meningitis in children in Kosovo. J ChildNeurol. 2012;27(1):46–50.

16. Nickerson JW, A Attaran, BD Westerberg, et al. Fatal bacterial meningitis possibly associated with substandardceftriaxone–Uganda, 2013. MMWR MorbMortal WklyRep. 2016;64(50–51):1375–1377

17. Chang JB, Wu H, Wang H, et al. Prevalence and antibiotic resistance of bacteria isolated from the cerebrospinalfluidofneurosurgicalpatientsatPekingUnionMedicalCollegeHospital. AntimicrobR esist InfectControl. 2018.

18. Children Hospital of king Pediatrics volume 139, king Daughters, Pediatrics volume 139, number ; e20163268.5, May 2017 https://doi.org/10.1542/peds.2016-3268

19. John E, Bennett MD, in Mandell, Douglas, and BennettsPrinciplesandpracticeofInfectious Diseases 2020. ISBN: 978-0-4430-6839-3

20. Spects AM, Bolijin R, Vanttor R. Global Etiology of bacterial meningitis; a systemcic review of metanalysis. PLOS one 2018;13: e0198772

21. Borer A, Saidel-Odes L, Riesenberg K, et al. Attributablemortalityrate for carbapenem resistant Klebsiellapneumoniaebacteremia. Infect 21.Control HospEpidemiol. 2009.

22. Lu C-H, Chang W-N, Chang H-W. Adultbacterialmeningitisinsouthern Taiwan:epidemiologictrendandprognosticfactors. JNeurolsci.24.2000.

23. Syeda Ayesha Ali, Muhammad Kamran Taj, Syeda Hafsa Ali, InfectionandDrugResistance 2021:145107–512028.

24. Cdc. How Antibiotic Resistance Happens. Centersfor Disease ControlandPrevention;March20,2019.Availablefrom:https://www.cdc.gov/drugresistance/about/how resistance-happens.html. Accessed November26,2021. doi:10.1001/jama.2021.2437

25. Bilal H,Khan MN, Rehman T, Hameed MF, Yang X. Antibio 9tic resistanceinPakistan:asystematicreviewofpastdecade. BMC InfectDis. 2021.

26. Syeda Ayesha Ali, Muhammad Kamran Taj, Syeda Hafsa Ali Infectionand Drug Resistance 2021. doi 10.2147/IDR.S339231

27. CDC. howantibioticresistancehappens. centersfordisease controlandprevention;march 20, 2019. available from: https://www.cdc.gov/drugresistance/about/how-resistance-happens.html., accessed november 26, 2021. doi:10.1001/jama.2021.2437

